# Systematic review of contemporary improvement interventions on care coordination, discharge support and transitional care from the patient experience perspective

**DOI:** 10.1101/2024.02.08.24302505

**Authors:** Tiago S Jesus, Brocha Z. Stern, Dongwook Lee, Manrui Zhang, Jan Struhar, Allen W. Heinemann, Neil Jordan, Anne Deutsch

## Abstract

**Aim:** To synthesize the impact of improvement interventions related to care coordination and discharge support on patient experience measures.

**Method:** Systematic review. Searches were completed in six scientific databases, five specialty journals and through snowballing. Eligibility included studies in English (2015-2022) on improving care coordination, discharge support, or transitional care assessed by standardized patient experience measures as a primary outcome. Two independent reviewers made eligibility decisions and performed quality appraisals.

**Results:** Of 1087 papers initially screened, 15 were finally included. Seven studies (three randomized controlled trials [RCTs]) focused on care coordination activities and used enhanced supports (e.g., improvement coaching; tailoring for vulnerable populations) for Patient-Centered Medical Homes or other primary care sites. Effectiveness was mixed or neutral relative to standard supports or models of care. Eight studies (three RCTs) focused on enhanced discharge support, including patient education (e.g., “teach back” method) and telephone follow-up or transitional support; mixed or neutral results on the patient experience were also found and with more substantive risks of bias.

**Conclusion:** Enhanced supports for improving care coordination, discharge education, and post-discharge follow-up had mixed or neutral effectiveness for improving the patient experience with care, compared to standard or simpler improvement approaches. Studies on the improvement of patient experiences, especially for enhanced patient discharge, need further strengthening such as including the patient perspective for its co-development.

## Background

The *patient experience* with care is an integral component of healthcare quality and a construct used to assess the *person-centeredness* of healthcare delivery, informing quality-improvement (QI) activities.^1–3^ Specifically, *patient experience* refers to how patients have experienced valuable aspects of healthcare delivery, such as physician communication, involvement in decision-making, getting timely appointments, or supports to care coordination and discharge planning in the transitions across settings to address care fragmentation issues.^1^

As synthesized in several systematic reviews, better patient experiences with care have been associated with improved treatment adherence and to improved patient and health system outcomes (e.g., reduced healthcare utilization or malpractice litigation).^4–7^ For example, a recent systematic review found that better scores on standardized patient experience measures were associated with greater self-reported physical and mental health, lower frequency and length of hospitalizations, and fewer emergency room visits.^4^ Furthermore, improving the patient experience of care has been recognized as a QI aim in itself, under the goal of improving the person-centeredness of care.^8 9^

Many health systems increasingly reimburse providers for the value of care, including patient experience scores. The US Agency for Healthcare Research and Quality (AHRQ) has developed the Consumer Assessment of Healthcare Providers and Systems (CAHPS) surveys and quality measures, which have been widely used to track, publicly report, and compare providers’ patient experience performance.^10^ Within selected pay-for-performance programs, the US Centers for Medicare & Medicaid Services uses CAHPS scores, reflecting patient experience of care, when computing payment incentives.^11^ Several other countries require providers to monitor and improve patient experience.^12 13^ Overall, healthcare systems, organizations, and frontline practitioners are increasingly subject to quality mandates, value-based incentives, or market pressures (e.g., customer loyalty) to develop systematic activities to improve their patient experience performance,^14^ including for care coordination and discharge support.^15 16^

Reducing care fragmentation, improving continuity of care, and preventing medication errors or rehospitalizations can be achieved by interventions such as supported discharge, case management programs, transitional care services, use of care coordinators as well as models of care such as patient-centered medical homes (PCMHs). Systematic reviews have addressed the effectiveness of these health service interventions, and many of these have been shown to be effective for a set of health, quality of life, and health system outcomes (e.g., reduced rehospitalizations).^17–20^ For example, interventions to facilitate the transition from hospital to home when started in the hospital and continued in the community through telephone or other telehealth follow-up or scheduled home visits can be effective in reducing hospital readmission, especially when tailored to each patient.^18 19 21^

However, we found no systematic review focused on the impact that care coordination, discharge support, or transitional care activities have on the patient’s experience with care. A recent overview of systematic reviews for care coordination interventions found patient experiences were rarely addressed or synthesized as outcomes of these interventions.^22^ Here, we aim to synthesis the contemporary literature (2015-2022) on the impact of care coordination, discharge support, and transitional care activities on patient experience as assessed by formal patient-reported experience measures.

## Methods

### Design

We registered (PROSPERO: CRD42022358337) a systematic review protocol that focused on synthesizing the English-language contemporary evidence (2015-2022) addressing health service interventions to improve the patient-reported experience with care. Here, we focus on the impact of interventions related to enhanced care coordination, discharge support, or transitional care activities. We use the Preferred Reporting Items for Systematic Reviews and Meta-Analyses (PIRSMA) guidance for the review report.

### Eligibility

We included controlled trials, longitudinal observational studies with controls, and pre-post designs with >30 participants. We excluded qualitative, cross-national studies, or any study not assessing the impact on patient-reported experience measures. Studies needed to have full texts available, focus on improving the patient experience (i.e., one of no more than two primary outcomes measures) and report inferential statistics (e.g., p-values) about the impact of the intervention on a patient experience measure.

For patient experience outcomes, we included studies that used standardized, quantitative patient experience assessments (e.g., validated surveys, their items or composite domains, or surveys that were externally collected and routinely used across providers, including for value-based reimbursement). We excluded studies that assessed patient experiences only with non-standardized, qualitative, or other non-validated instruments. Measures that were not patient-reported (e.g., observational) were excluded. We did not synthesize the impact of the interventions on measures other than patient-reported experience with care.

For participants, we included health systems, organizations, providers, networks, settings, or service units, including any health professionals or staff. We excluded health service interventions exclusively delivered by students or clinicians-in-training. Those providing patient experience feedback could be the patient, family/informal caregivers, or proxy respondents.

For context, we had no geographic restriction, but we only included English-language articles published in 2015 or thereafter, to reflect contemporary interventions responsive to the more recent focus, mandate, or incentives for improving the patient experience of care. For example, value-based reimbursements that included patient experience scores did not emerge in the US until 2013.^23^

For this review, we only included health-service improvement interventions focused on *care coordination* (i.e., systematic organizational, service line, or service unit strategies to address fragmentation of care delivery and enhance continuity of care^22^), *discharge support* (i.e., patient education in preparation for discharge and/or post-discharge education or follow-up support either in the environment or by telephone or other telehealth/web-based mechanisms^18^), and *transitional care* (i.e., subset of intermediate, time-limited set of actions to ensure the coordination or continuity of care as patients transfer between different settings or levels of care^24^), which often involves a single point of contact to optimize service access, communication, and coordination.^25^

### Search

We searched six scientific databases (PubMed/MEDLINE, CINAHL, EconLit, PsycINFO, DOAJ, and Scopus) using a combination of free-text words with indexed terms that reflect our eligibility criteria. We restricted the search for English language and publications after January 2015. **Supplementary Appendix 1** shows the detailed searches for each database.

We also conducted targeted searches within the *Patient Experience Journal*, *Journal of Patient Experience*, *Medical Care*, and *Health Expectations*. *NEJM Catalyst Innovations in Care Delivery* was searched since January 2020, when the journal became peer reviewed. These were journals with frequent papers identified in the initial database searches. Finally, we conducted snowballing strategies (e.g., citation-tracking, similar publications in PubMed) using the final included articles and reference lists of similar systematic reviews, including those identified through the searches.

### Selection

Two independent reviewers (TJ first reviewer; DL, JS, or MZ for the second reviewer role) conducted the title-and-abstract screenings and then the full text reviews against the eligibility criteria. Reviewer consensus was achieved on the final inclusion within a round of discussion, with no need for third reviewer involvement.

### Data Extraction

The research team built a data extraction form that they used to chart data such as study characteristics and context, patient experience measures and metrics, interventions, analytic approaches, and results pertaining to patient-reported experience with care. The data were charted initially by the first author (TJ) and verified by a second author (BS), with subsequent rounds for any flagged extractions until they reached consensus. **Supplementary Appendix 2** provides detailed, consensus-based data extractions for each included study.

### Quality Assessment

The quality assessment was performed independently by TJ and BS. The Cochrane-suggested risk of bias criteria for Effective Practice and Organization of Care (EPOC) reviews were used for the controlled trials, while the National Heart, Lung, and Blood Institute’s quality assessment tools were used for the pre-post studies and observational studies that assessed or compared the effects of health service interventions.

As planned in the review protocol, we did not grade the strength of the evidence across the research studies and their design. Covering a range of health service or QI interventions, contexts, assessments, and study designs, our review is configurative (not aggregative) in nature. We tabulated the consensus-based appraised risks of bias for each study’s context and design, based on the respective checklist-based methodological assessment; those detailed assessments are in **Supplementary Appendix 3**.

### Analytic plan

We tabulated and synthesized the results per two major inclusion categories: 1) care coordination and 2) discharge support and follow up. The latter included transitional care because the included studies that had this component were in relation to discharge support. Within each category, we tabulated the methods and appraised the risk of bias for each study, and then synthesized the interventions and their findings, ordered by study design (starting with RCTs) and publication date within the same design. Given the configurative nature of the review and the heterogeneity of interventions and study contexts, we did not conduct a meta-analysis.

## Results

Figure 1 provides the PRISMA flowchart.

**Figure 1:**
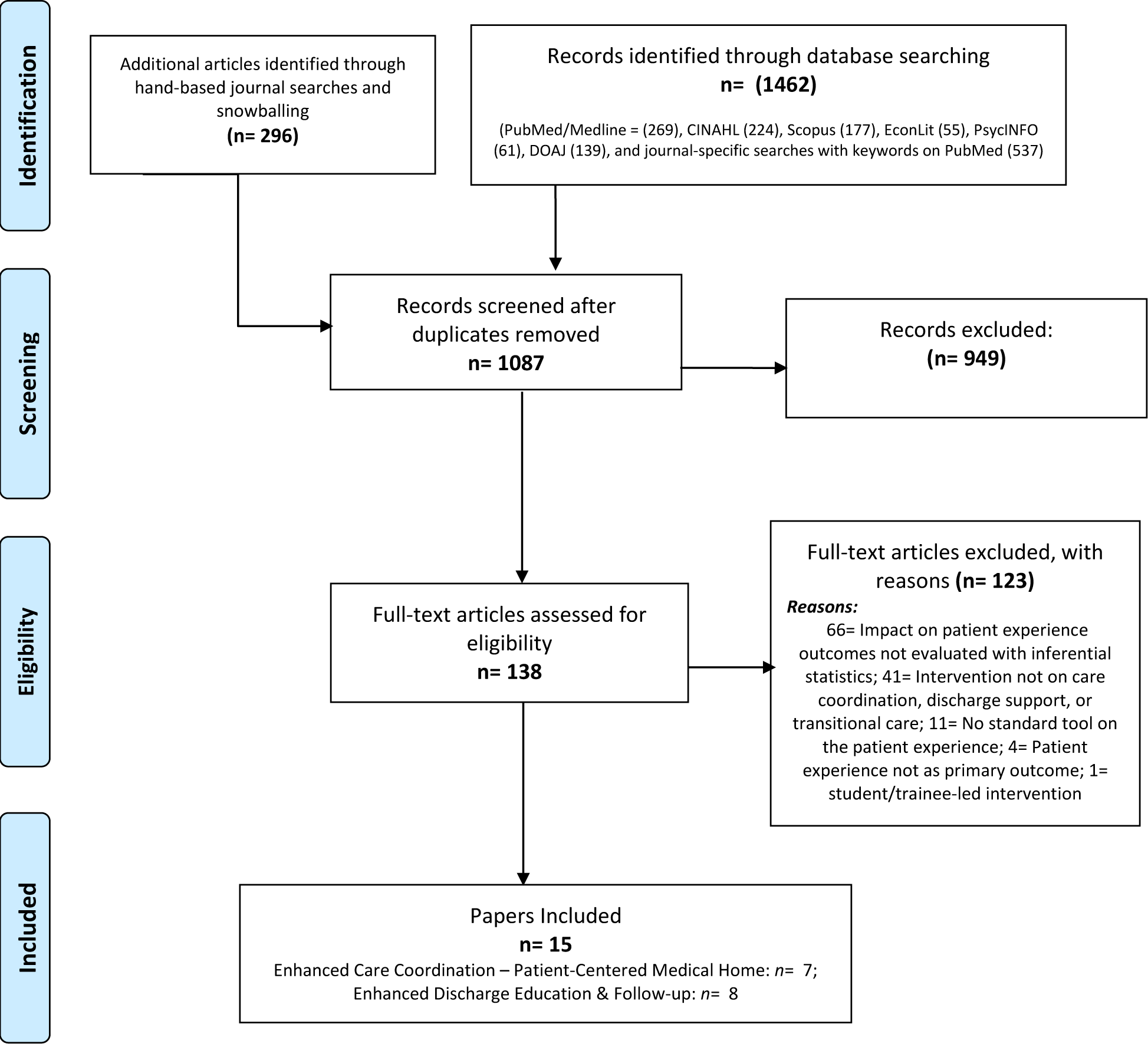
PRISMA flowchart of the systematic review.

Among the 1087 unique papers screened, 138 full texts were assessed for eligibility and 15 were included.^15 16 26–38^ The absence of impact assessment with inferential statistics was the main reason for exclusion at the full-text assessment. Among papers included, seven focused on enhanced care coordination supports (e.g., over and beyond models of care based on a PCMH), whereas eight focused on enhanced discharge supports, specifically for enhanced patient education, reengineered discharge, telephone follow-up including for transitional care, or a mix of those interventions.

a) Enhanced care coordination supports – primary care level (*n*= 7)

**Table 1** summarizes each study’s methods (e.g., design, context) and risks of bias; **Table 2** summarizes the intervention and study findings.

**Table 1:**
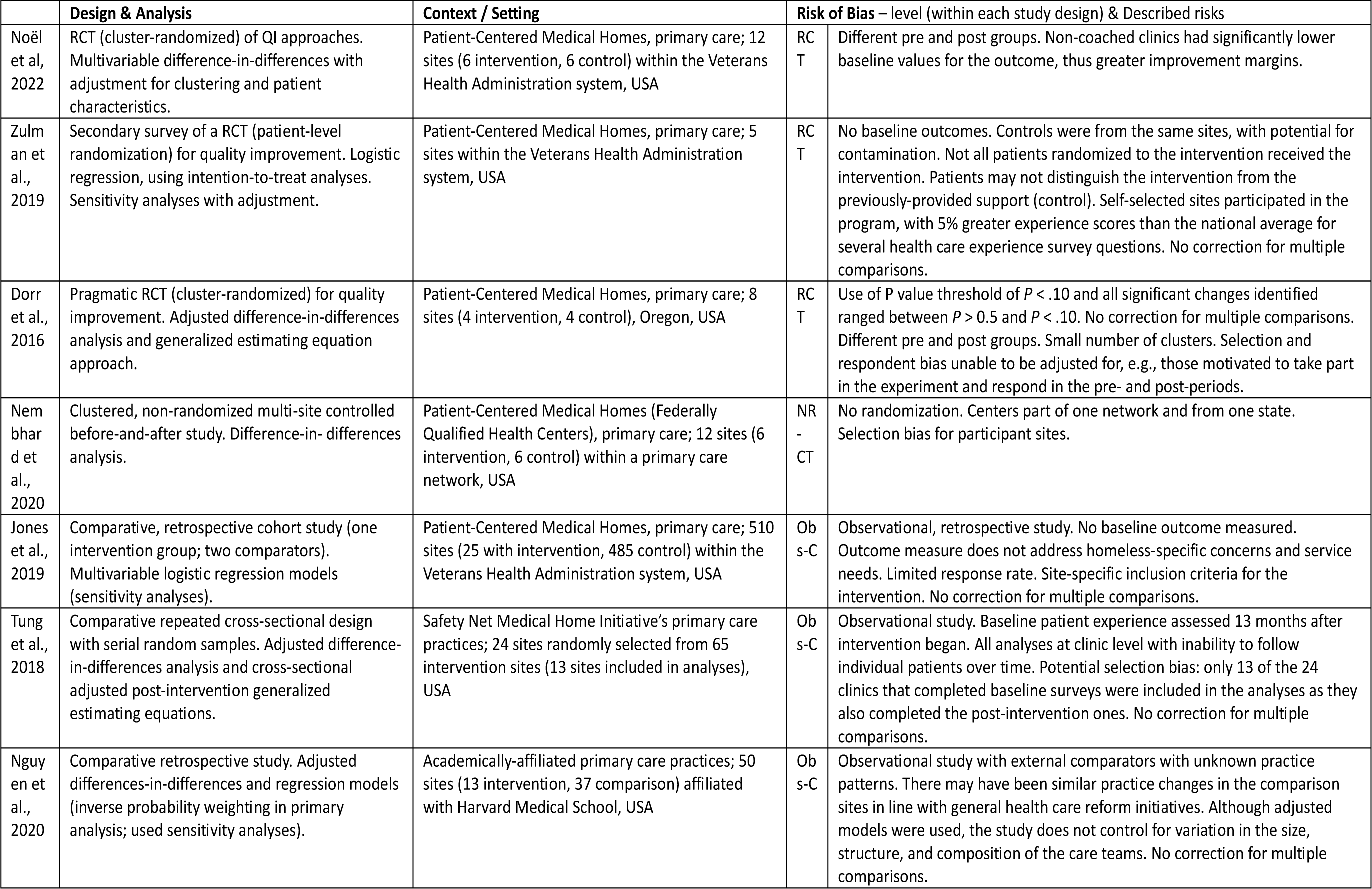
Summary of study methods and risk-of-bias assessment: Enhanced care coordination supported for Patient-Centered Medical Homes or other primary care practices. Legend: RCT: Randomized Controlled Trial; NR-CT: Non-Randomized Controlled Trial; Obs-C: Observational, comparative study. QI: Quality Improvement

**Table 2:**
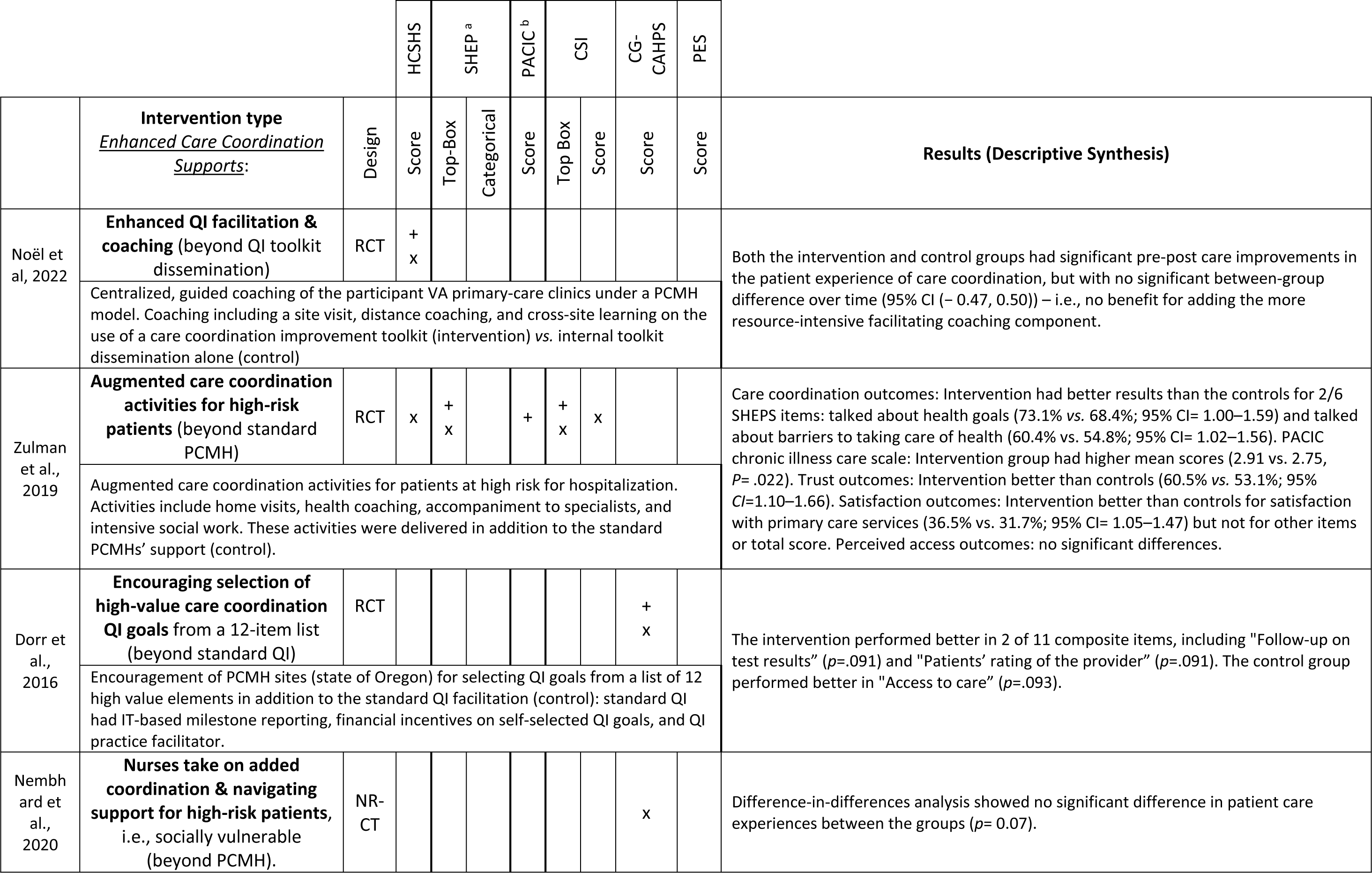
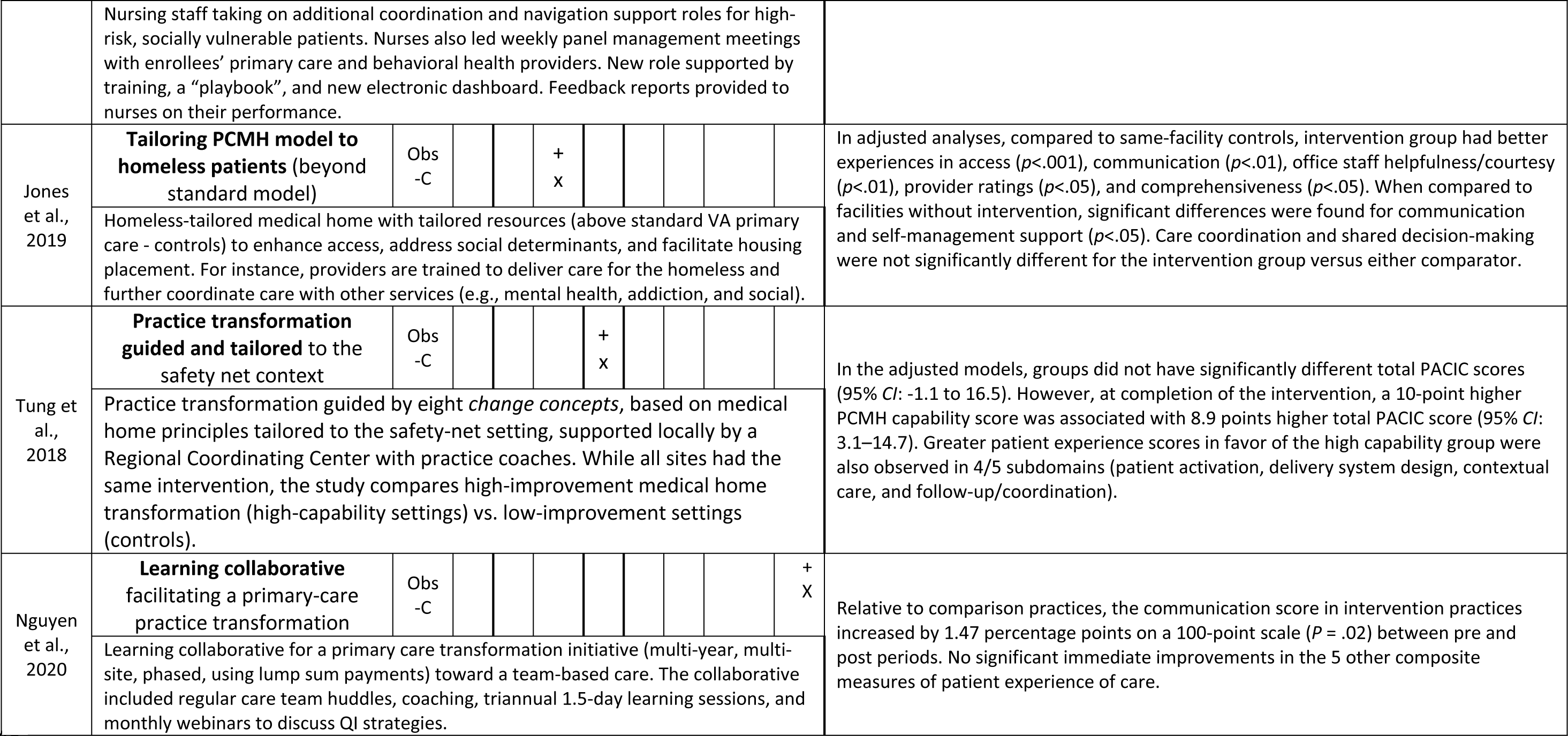
Enhanced care coordination for Patient-Centered Medical Homes (PCMHs) or other primary care practices evaluated from the patient experience perspective as a primary outcome. Keys: Results per measure & metric: + significant positive impact; x non-significant impact. Legend: HCSHS: Health Care (System) Hassles Scale; SHEP: VA’s Survey of Healthcare Experiences of Patients (adapted from CAHPS); PACIC: Patient Assessment of Chronic Illness Care; CSI: Consumer Satisfaction Index; CG-CAHPS: Consumer Assessment of Healthcare Providers and System Clinician & Group Survey; PES: Patient Experience Survey; RCT: Randomized Controlled Trial; NR-CT: Non-Randomized Controlled Trials; PPT: Pre-Post Test study; Obs-C: Observational, comparative study. QI: Quality Improvement. VA: Veterans Administration. IT: Information Technology. ^a^ Jones et al. specifically reported use of the PCMH-SHEPS. ^b^ Tung et al. reported use of a modified version of the PACIC.

Context-wise, all studies were multi-site PCMHs or other primary care practices, all from a network or integrated health system in the US: Veterans’ Health Administration (VHA),^32–34^ a statewide PCMH network,^35^ Federally Qualified Health Centers,^36^ a Safety Net Medical Homes’ Initiative,^38^ or academically-affiliated primary care practices.^37^

Three studies were RCTs for facilitating site-based improvement projects. A recent RCT at the VHA, with the fewest appraised risks of bias (Table 1), used QI facilitation (e.g., coaching) on the site’s use of a care-coordination improvement toolkit – beyond its internal dissemination (control); both groups improved, with no significant benefit for the more resource-intensive arm.^34^

Another RCT focused on the impact of augmented care coordination for patients with high risk of hospitalization (beyond standard PCMH; control group) and found improvements in 2 of 6 care coordination items.^33^ The remaining RCT encouraged the selection of high-value care-coordination QI goals (beyond standard QI facilitation: control group) and found mixed results.^35^ These RCTs had some appraised risks of bias (e.g., potential contamination;^33^ selection bias^35^).

Among other study types, one non-randomized trial focused on nurses taking on additional care coordination and navigation support for socially vulnerable patients, beyond a PCMH model (control), and found close but no significant differences between groups (*p*= 0.07).^36^ Finally, three observational studies (tailoring PCMH model to homeless patients;^32^ practice transformation tailored to the safety nets;^38^ learning collaborative^37^) showed mixed results and had substantive risks of bias (Table 1).

b) Enhanced discharge or follow-up supports - after hospital or surgical care (*n*= 8)

**Table 3** summarizes the methods and **Table 4** the interventions and results for the studies on enhanced discharge support, including patient education or follow-up. All studies were based in the US.

**Table 3:** Summary of study methods and risk-of-bias assessment: discharge education and follow-up or transitional support from an inpatient or surgical setting. Legend: RCT: Randomized Controlled Trial; NR-CT: Non-Randomized Controlled Trial; PPT: Pre-Post Test study; Obs-C: Observational, comparative study.

|  | Design & Analysis | Setting | Risk of Bias – level (within each study design) & Described risks |  |
| --- | --- | --- | --- | --- |
| van Eck et al., 2018 | Single-site RCT (patient-level randomization) with posttest measure only. T test and analysis of variance on covariates. | Private practice orthopedic clinic, outpatient and ambulatory surgery; Kerlan Jobe Orthopaedic Clinic, California, USA | RCT | Single site with 2 surgeons. Patients not blinded to group allocation. No baseline outcomes. No correction for multiple comparisons. Limited follow-up time (only two weeks of post-assessment). |
| Griffey et al., 2015 | Single-center RCT (patient-level randomization) with intention-to-treat analysis. Multivariable ordinal logistic regression. | Hospital, emergency department; Barnes Jewish Hospital, St. Louis, Missouri, USA | RCT | No baseline outcomes assessed. Single site. Convenience sampling prior to randomization. About 40% loss to follow-up. No random sequence generation for group assignment or allocation concealment. Contamination may have occurred. No correction for multiple comparisons. |
| Chan et al., 2015 | RCT (patient-level randomization stratified by language). $\chi^2$ test on the top-box scores. Sensitivity and subgroup analyses. | Safety-net hospital, various units (internal medicine, family medicine, cardiology, neurology); San Francisco General Hospital and Trauma Center, California, USA | RCT | No baseline outcomes assessed. Single site. Only one month of post-intervention data. Medical teams not blinded to presence of intervention, and possible contamination across nurses within the hospital. No correction for multiple comparisons. |
| Centrella-Nigro & Alexander, 2017 | Nonrandomized trial, two units of one hospital. Independent T tests. | Community Magnet-designated hospital, medical units (intervention unit mainly neurological patients, control unit mainly genitourinary and infectious diseases); Northern New Jersey, USA | NR-CT | No randomization. Clinical differences in units. Difference in baseline outcomes not adjusted for. Baseline characteristics not assessed or adjusted for. Potential for contamination because all units were in a single hospital. No correction for multiple comparisons. Lack of transparent reporting of participant numbers pre- and post-intervention. No combined analysis of between-group and between-time. |
| Cancino et al., 2017 | Nonrandomized posttest design, two control groups with standard discharge (one in same unit and one in different unit). $\chi^2$ test on the top-box scores. | Safety-net hospital, adult inpatient family medicine (adult general medical units as one comparator); Boston Medical Center, Massachusetts, USA | NR-CT | No randomization. No baseline characteristics or outcomes data. No adjustment for covariates or correction for multiple comparisons. Low response rate. Possibility for contamination as some controls were within the same unit. Nurses selected patients (potential selection bias). Results not transferable to sites with higher baseline scores. |
| Schreiter et al., 2021 | Historical cohort of matched controls at patient level. Wilcoxon rank-sum for scores and Fisher exact test for % of top-box scores. | Academic hospital, transition from surgery; University of Wisconsin Hospital, Wisconsin, USA | Obs-C | No randomization. Historical controls even though matched for some clinical indicators. Single site. No baseline outcomes assessed. Unadjusted analyses for outcome of patient experience. No correction for multiple comparisons. |
| Thum et al., 2022 | Pre- and post-test, one organization with multiple units. $\chi^2$ on top-box scores and Kruskal-Wallis for the percentile rank change. | Academic tertiary care center and its affiliated community hospital; Thomas Jefferson University Hospital and affiliate, USA | PPT | No control group. Eligibility criteria not clearly described. Single pretest (although long preintervention period). No establishment of stable baseline through multiple measurement time points. No report on the numbers of surveys used for the analysis, only those mailed. No correction for multiple comparisons. |
| March et al., 2022 | Pre- and post- test, single-center retrospective review of a pilot program. $\chi^2$ test on the top-box score after T test on baseline characteristics. | Hospital, pharmacy; Methodist University Hospital, Memphis, Tennessee, USA | PPT | Retrospective, single-site pre-post-test analysis not adjusted for covariates. No establishment of stable baseline through multiple measurement time points. No correction for multiple comparisons. Intervention not delivered to all the eligible patients (business hours). No report on the number of patient experience surveys collected. Risks of selected findings: two different designs for two outcomes. |

**Table 4:**
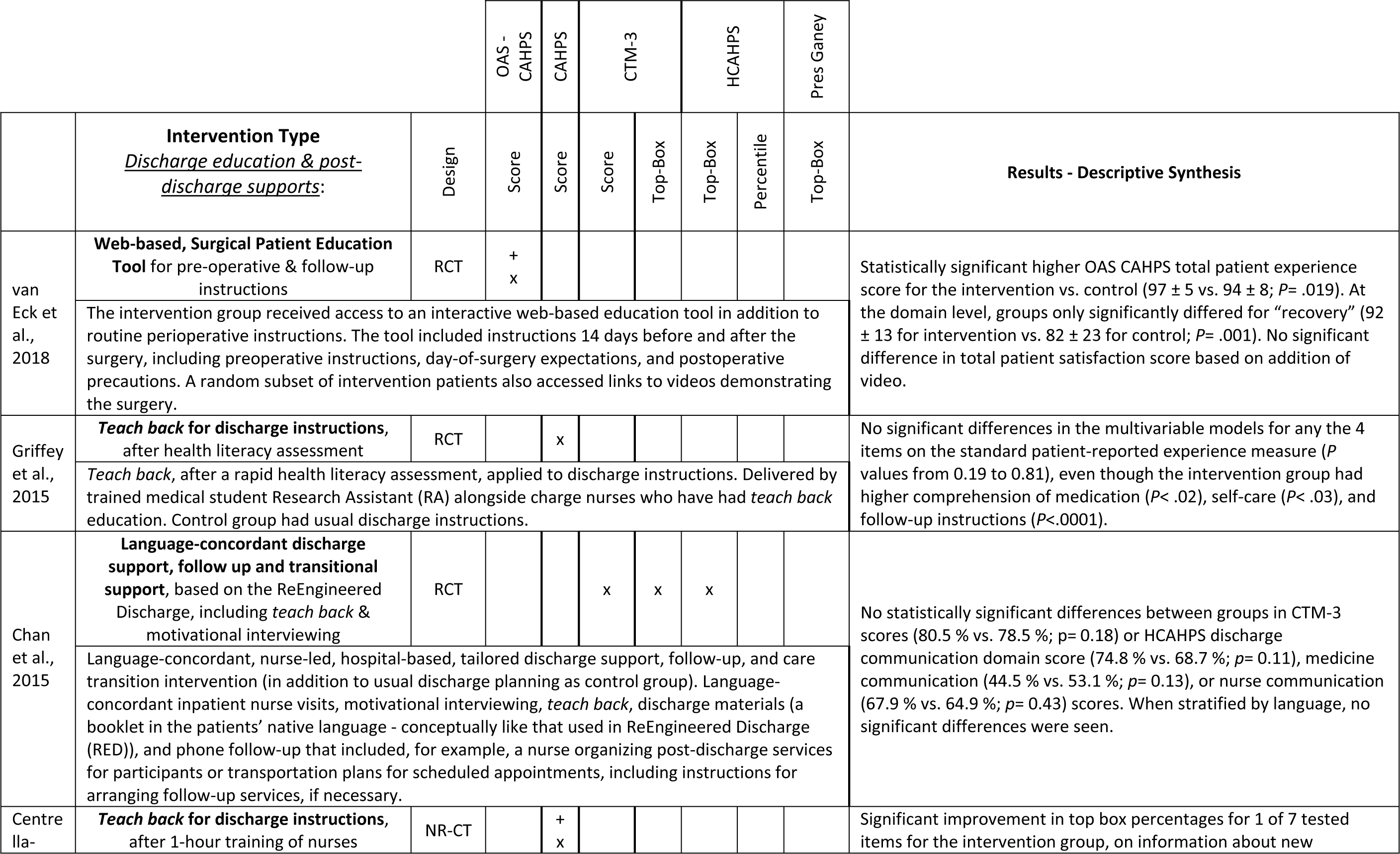
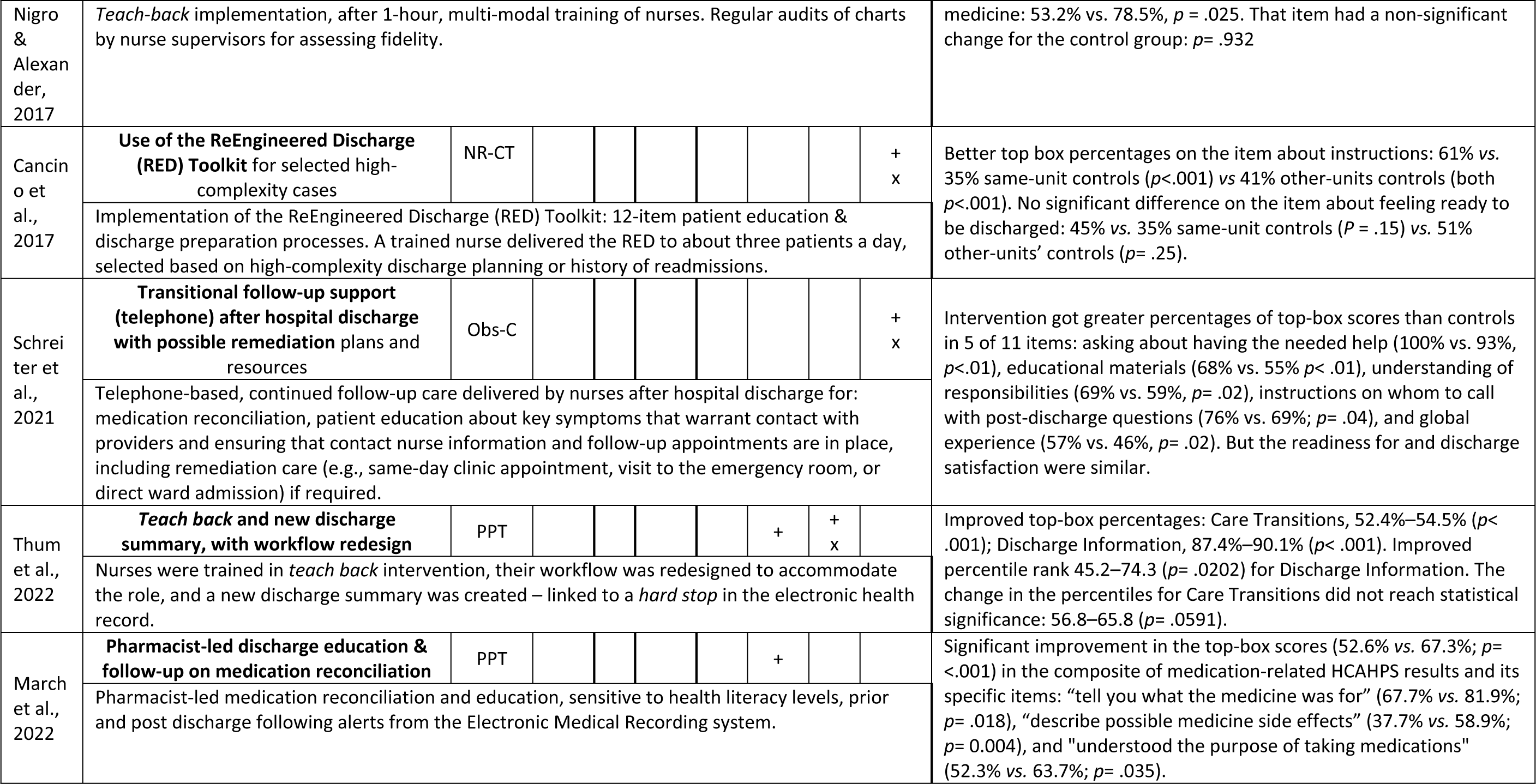
Discharge education and post-discharge follow-up (e.g., transitional care supports) after inpatient or surgical care evaluated from the patient experience perspective as a primary outcome. Legend: OAS-CAHPS: Outpatient and Ambulatory Surgery Consumer Assessment of Healthcare Providers and System; CAHPS: Consumer Assessment of Healthcare Providers and System; CTM-3: Care Transitions Measure; HCAHPS: Hospital Consumer Assessment of Healthcare Providers and Systems; RCT: Randomized Controlled Trial; NR-CT: Non-Randomized Controlled Trials; PPT: Pre-Post Test study; Obs-C: Observational, comparative study.

Among the three RCTs, two focused on patient discharge education using the *teach back* method, after a health literacy assessment^26^ or as part of a multi-modal language-concordant discharge support and telephone follow-up.^27^ None of the study interventions led to better experience results than standard care. The other RCT was on a web-based, surgical patient education tool for pre-operative & follow-up instructions, and found improvements in the overall experience score.^28^ However, several risks of bias were appraised in all the RCTs such single-center study,^26–28^ limited follow-up,^27 28^ or no correction for multiple comparisons (Table 3).^26-28^

Two non-randomized trials were found: one on the implementation of *ReEngineered Discharge* for three patients a day that nurses selected as complex cases,^16^ the other on *teach back* discharge education by nurses.^31^ Both achieved improvements in selected patient experience items (e.g., information provision). In turn, an observational study with historical controls focused on a telephone follow-up after hospital discharge with possible remediation plans or rescue resources (e.g., same-day clinic appointment);^15^ it achieved higher percentages of top-box scores (i.e., highest score on the given patient experience items) compared to controls for 5 of the 11 patient experience items. Several risks of bias applied to the three studies (Table 3), including risks of contamination and unadjusted analyses.

Finally, two pre-post studies were included, one using *teach back* and other discharge redesigns,^29^ the other a pharmacist-led discharge education and follow-up intervention.^30^ These provided either positive or mixed results (improvement in top box percentages but not in the percentile ranks with peer providers). In addition to the uncontrolled study design, these studies had multiple substantive risks of bias (Table 3), limiting the ability to draw conclusions.

## Discussion

This knowledge synthesis addresses a gap in the literature regarding the impact of care coordination or discharge support improvement on standardized patient experience measures. Health-service improvement activities, including enhanced supports or tailoring for vulnerable populations, achieved mixed to null improvements in the patient experience with the care coordination or discharge processes, compared to standard models of care or simpler quality-improvement supports. This was found by seven studies (including three multi-site RCTs) on enhanced QI support provided to PCHMs or other primary care practices. It was also found by eight studies on enhanced discharge support such as patient education, reengineered discharge, and/or telephone follow-up or transitional support. These latter studies, as a body of literature, had a greater risk of bias.

Several risks of bias were identified in the reviewed literature across the study designs, limiting the internal and external validity of the findings. The studies on enhanced discharge support were typically single center. Even the multi-site studies on enhanced care coordination were performed within a network or organization. Hence, it is unknown how the findings apply elsewhere. For instance, the study with the lowest appraised risks of bias found that the more resource-intensive type of QI support provided to PCMH units was not superior to the dissemination of the QI tool alone for driving improvements in patient experience of care coordination activities. But this finding directly applies to the VHA. The VHA has been subject to systematic QI programs, studies, or activities,^39 40^ which may have contributed to building the capacity for effective QI activities across their units as merely supported by an online QI tool, even without a centralized and more costly QI facilitation. The same may not apply to other healthcare organizations and development circumstances. Because context is a vital variable in QI science,^41^ future studies may consider generating evidence with validity across organizations and networks and their varied sets of circumstances. QI collaboratives or hubs, including those externally funded, might equitably foster the capacity to develop and assess QI activities across service delivery organizations, say within a jurisdiction within the US.^42^

Varied study designs were included here. While RCTs can produce the most solid evidence when well conducted, a wider range of designs were included to reflect real-world improvement activities and evaluations, including pragmatic constraints.^43^ Study limitations within each design were reflected in the appraised risks of bias, especially among the single-center discharge support studies. Moreover, studies’ delivery of the tested intervention was sometimes pragmatically constrained, too. For instance, a pharmacist-led patient education intervention was performed for patients discharged only during particular hours.^30^ Similarly, a reengineered discharge support program was capped at a number of patients per day as selected by nurses as “complex” cases.^16^ These limitations may affect the fidelity of the improvement interventions and accuracy of the results and may often reflect budget or other limitations (e.g., human resources, in-house improvement or research expertise) for the conduct or evaluation of QI activities.^43^ Percentile ranks (i.e., provider ranks in patient experience performance relative to peers) were used as an additional metric only in one included paper (with a pre-post design), showing mixed results while the percentage of top-box scores had positive results.^29^ Reporting results in more than one metric of a patient experience measure can be important, especially for percentile ranks in pre-post studies to partly overcome the lack of a control group or the use of historical ones. Overall, the inclusion of varied study designs, interventions, and patient experience measures and metrics may inform further improvement activities and its study, at varying levels of available resources. Across study designs, some common intervention approaches were found, such as enhanced supports for QI facilitation on care coordination^34 35^ or tailored approaches for at-risk populations (e.g., socially vulnerable, homeless),^32 33 36 38^ often showing mixed patient experience results relative to standard supports. In turn, discharge support approaches that used a *reengineered discharge* or *teach back* strategies were also common but with less favorable results when compared to studies and reviews on other outcomes, such as reducing readmissions.^44–46^ For example, one recent systematic review of *teach back* strategies applied to discharge education identified a 45% reduction in 30-day readmission.^46^ Here, studies using the *teach back* method had mixed results overall^26 27 29 31^ but neutral results in the RCTs,^26 27^ either as the main strategy^26^ or as part of a multi-modal strategy that also included a reengineered discharge, motivational interviewing, follow-up and transitional support, and overall language-concordant supports.^27^ When found to be effective compared to a control intervention, the *teach back* method was used in isolation and specifically improved the patient experience with receiving information about new medicines.^31^ For transitional care post discharge, which was rarely addressed in the body of literature reviewed, the multi-modal intervention that included this component found neutral results,^27^ whereas the intervention specifically focused on this component found positive results in five patient experience items.^30^ Further research could compare the cost-effectiveness of multi-modal and single-method improvement approaches as well as determine whether multi-modal interventions, due to their complexity, are delivered with fidelity to the main active ingredients of the intervention.^47 48^

To be impactful on the patient experience, it may be key for studies to assess how patient-centered (e.g., how attentive, respectful, responsive, with genuine interest) was the delivery of a *teach back*, telephonic transitional support, and other supportive patient-provider communications intervention. This might include how tailored the communication was to the individual patient^18^ to avoid the risks of being perceived as just another *box-ticking* exercise.^49^ Finally, in alignment with growing focus on coproduction and on experience-based codesign for health-service improvement activities,^50 51^ the patient experience with care can be further engaged all the way through from the intervention codesign to its evaluation. The literature reviewed here had no focus on engaging the patient perspective in the codesign of the improvement interventions.

All the included studies were conducted in the US, which may result from multiple factors. First, the CAHPS program in the US provides established, widely used standardized patient experience measures, which was a criterion for inclusion. Second, providers’ performance on CAHPS measures is publicly reported and included in some value-based purchasing programs,^11^ which may have driven the interest of providers to develop and report these improvement. Third, CAHPS has been a primary quality target of a “person-centered” medical home, with the US National Committee for Quality Assurance recommending administration of the CAHPS PCMH survey for PCMH transformation, and a related study found that CAHPS surveys were considered actionable for PCMH transformation.^52^ Fourth, the VHA in the US has its own patient experience measurement system, which was used a standardized assessment measure in studies externally reported and included here.^33 34^ Fifth, improvement activities otherwise fitting the study eligibility criteria from other countries may have been conducted but not reported in English or through peer-reviewed journals.

This review has several limitations. First, it excluded articles published in languages other than English or in the grey literature, which affects representativeness but reflects peer-reviewed knowledge readily available to an international audience. Second, representativeness is also affected by the exclusion of articles published before 2015, which allowed for a synthesis of contemporary approaches in an ever-changing healthcare delivery landscape. Third, for the data extraction, the second reviewer performed only confirmation tasks while two independent reviewer roles were used for all other tasks. Fourth, we did not perform a formal grading of the evidence or of the risks of bias within or across study types, within heterogenic health service delivery contexts, interventions, and study methods. Fifth, we did not include studies that used patient experience as a secondary study outcome. While including these studies may have added to the pool of evidence, it would have risked including studies in which improving the patient experience of care was not a primary intention.

## Conclusion

Enhanced support for improving care coordination and discharge support had mixed or neutral results for improving the patient experience with care beyond standard care or simpler supports. Substantial risk of bias and lack of comparison between improvement approaches impede firmer conclusions, especially for the studies on enhanced discharge support, which were also typically single center. The multi-site studies on enhanced care coordination were also limited by being performed within an integrated delivery system, network, or organization, limiting the generalizability of the results across organizational and service delivery contexts. Improving the person-centeredness of care may require strengthening the capacity to fund, design, conduct, and evaluate improvement studies focused on patient experience across providers and their networks. Improving the patient experience with care coordination and discharge may require the strengthening of the improvement studies, and further engaging the patient perspective from the intervention development to its evaluation.

## Data Availability

The data are provided as supplementary materials.

